# Outdoor PM_2.5_ Concentration and Rate of Change in COVID-19 Infection in Provincial Capital Cities in China

**DOI:** 10.1101/2020.05.19.20106484

**Authors:** Yang Han, Jacqueline CK Lam, Victor OK Li, Jon Crowcroft, Jinqi Fu, Jocelyn Downey, Illana Gozes

## Abstract

Motivated by earlier findings that exposure to daily outdoor PM_2.5_ (P) may increase the risk of influenza infection, our study examines if immediate exposure to outdoor P will modify the rate of change in the daily number of COVID-19 infections (R), for (1) the high infection provincial capital cities in China and (2) Wuhan, China, using regression modelling. A multiple linear regression model was constructed to model the statistical relationship between P and R in China and in Wuhan, from 1 January to 20 March 2020. We carefully accounted for potential key confounders and addressed collinearity. The causal relationship between P and R, and the interaction effect between key variables were investigated. A causal relationship between P and R across the high infection provincial capital cities in China was established via matching. A higher P resulted in a higher R in China. A 10 µg/m^3^ increase in P gave a 1.5% increase in R (*p* < 0.001). An interaction analysis between P and absolute humidity (AH) showed a statistically significant negative relationship between P × AH and R (*p* < 0.05). When AH was $ 5.8 g/m^3^, a higher P and AH gave a higher R. In contrast, when AH ≥ 5.8 g/m^3^, the effect of a higher P was counteracted by the effect of a higher AH, resulting in a lower R. Given that P can exacerbate R, we recommend the installation of air purifiers and better air ventilation to reduce the effect of P on R. Further, given the increasing discussions/observations that COVID-19 can be airborne, we highly recommend the wearing of surgical masks to keep one from contracting COVID-19 via the viral-particulate transmission pathway.

## 1. Introduction

COVID-19 was first reported in Wuhan, China in December 2019. Since the outbreak, COVID-19 has resulted in more than 116-million infections and 2-million deaths globally, with more than 100,000 reported cases in China.

Recent COVID-19 studies have investigated if demography (D), co-morbidity (CM), meteorology, and lockdown generate any significant statistical effects on viral infection.^1–3^ Evidence so far implies that low temperature and humidity are associated with COVID-19 transmission.^2^ Such observation is consistent with earlier epidemiological studies implicating that air pollutants (PM_10_ as the primary pollutant) and meteorologies are associated with SARS/MERS infection.^4,5^ Furthermore, influenza studies have suggested that exposures to PM_2.5_ (P) with and without interacting with meteorology may increase the risks of influenza infection.^6^ In the US and Europe, long-term exposures to P and NO_2_ are linked to COVID-19 mortality.^7,8^ Air pollution is considered to heighten the severity of COVID-19 infection, given that pollutants, such as P, may increase the risk of Vitamin-D deficiency and decrease immunity.^9^ To date, increasing evidence in China, Italy, and other countries has shown that air pollution is a significant contributor to COVID-19 infection.^10–12^ COVID-19 studies conducted in China conclude that P, NO_2_, and O_3_ are associated with the number of COVID-19 infections,^13^ with significant interaction effect between air quality index (AQI) and rising temperature identified.^14^ However, such studies have failed to fully account for the change in testing capacity and the inconsistency in COVID-19 case definition, as well as the confounding effects of D and CM. A more sophisticated and rigorous study conducted in Italy, utilizing doubling time derived from a fitted epidemic curve to measure COVID-19 transmission, and after removing potential confounders and noises, has concluded that P alone does not facilitate COVID-19 transmission across the most affected regions.^15^ However, a study conducted in UK has suggested a positive relationship between P and COVID-19 infection cases, after controlling for D and CM confounders, including, population density, age, sex, diabetes, smoking status, and cancer.^16^ The above suggests whether short-term P exposure affects COVID-19 infection in China remain inconclusive, given the existence of noises and irregularities underlying the epidemic trends, the lack of control of confounders to P exposure, and the lack of sophisticated models to control these data challenges. More rigorous statistical modelling and control methodologies are needed to reduce (1) the noises underlying the epidemic trends due to the lack of testing capacity and redefinition of confirmed cases, (2) the confounding biases that affect the causal link between P and COVID-19 infection, and (3) the collinearities across different meteorology, D, and CM variables.

In this study, we will examine the immediate effect of outdoor P on the rate of change in the daily number of COVID-19 confirmed infections (R), across 18 high infection provincial capital cities in China, while seeking to address inadequacies in official case reporting due to the lack of testing capacity and inconsistencies in case definition, and taking into account as many confounders as possible, including D, CM, meteorology, net move-in mobility (NM), time lag due to the incubation period, trends over time (T), and day-of-the-week (DOW) to reflect the recurrent weekly effect (see Table S5 in Appendix p 3-5 for definitions on the variables).

Outdoor P is chosen as the focus of our study given the assumption that R may be increased due to the potential deposition of viral droplets on P.^17^ A recent rigorous study on COVID-19 aerodynamics ascertained that viral aerosol droplets 0.25-1 µm in size can remain suspended in air.^18^ When such viral droplets are combined with suspending particles, P, they can travel greater distances, remain viable in the air for hours, and be inhaled deeply into the lungs, thus increasing the potential of airborne viral infection.^19^

Our study sheds new light on the effect of P in an outdoor environment, the interaction effect between P and absolute humidity (AH), and the effect of NM (lockdown), on R (the dependent variable). Our work adds weights to the recent discussions/observations that COVID-19 droplets are airborne^18,20^, can suspend in the air and combine with the particulates, promoting infection via the airborne transmission pathway.^21^

Given that P can exacerbate R, we recommend the installation of air purifiers and improving air ventilation to reduce the effect of P on R. Further, given the increasing discussions/observations that COVID-19 can be airborne, the wearing of surgical masks are highly recommended to protect one from contracting COVID-19 via the viral-particulate transmission pathway.

## 2. Results

### 2.1 Descriptive Statistics and Data Adjustments

We collected data, including the number of confirmed COVID-19 cases, PM_2.5_ pollution, meteorology, mobility, demographics, and co-morbidities, in 31 provincial capital cities in China, covering the period from 1 January to 20 March 2020 (see Section 4.1 for more details). The spatial distribution of COVID-19 infection in the 31 provincial capital cities in China is shown in Figure 1(a). The collected COVID-19 infection data was pre-processed (see Section 4.2 for more details). 13 cities were removed due to small sample size (i.e., less than 50 confirmed cases in total). The remaining 18 cities (including Wuhan) were considered the COVID-prone provincial capital cities and were kept for further analysis. Due to the potential delays in case reporting and redefinition of confirmed cases, the COVID-19 infection data in the high infection provincial capital cities were adjusted by a moving average interpolation method and an outlier removal procedure, with the aim to reduce the short-term fluctuations in the reporting of COVID-19 confirmed cases and to recover the underlying COVID-19 epidemic trends. Taking Wuhan, the epicenter of COVID-19 in China during the study period, as an example, Figures 2(a) and 2(b) demonstrate the epidemic trend of COVID-19 infection in Wuhan before and after the data adjustments. Further, the adjusted daily confirmed cases were used to calculate R, a metric that measures the relative change of COVID-19 infection. By using R, even if the number of reported infections might deviate, the relative change in infection could still be accounted for, provided that the adjusted data can reflect the underlying trends of COVID-19 infection. Figure 2(d) shows the adjusted distribution of R in the high infection provincial capital cities in China for the statistical analysis.

**Figure 1.**
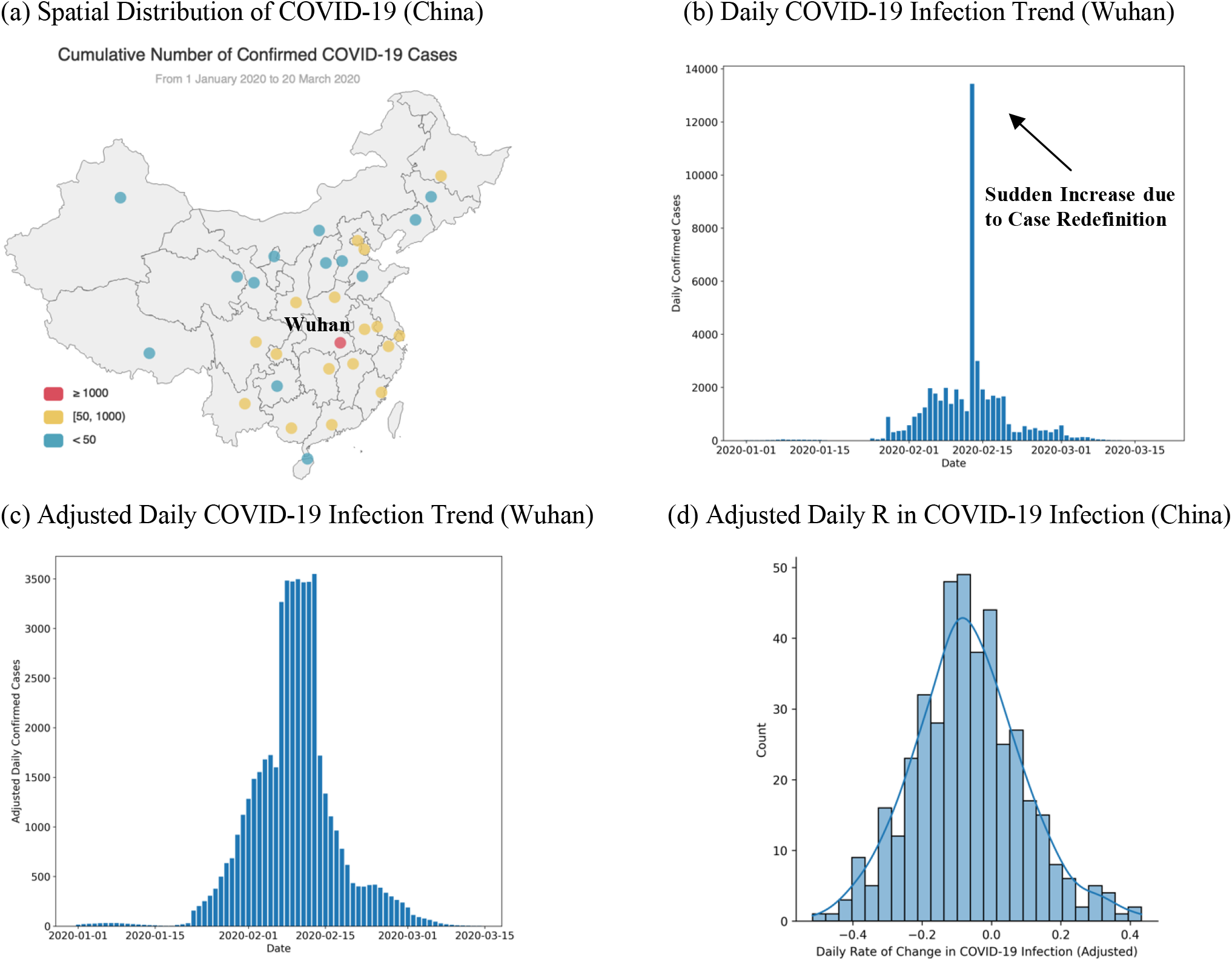
(a) the distribution of the cumulative number of confirmed COVID-19 cases in the provincial capital cities in China (including Wuhan), (b) the daily number of COVID-19 infection in Wuhan, (c) the adjusted daily number of COVID-19 infection in Wuhan, and (d) the adjusted daily R in COVID-19 infection in the high infection provincial capital cities in China (including Wuhan), during 1 January 2020 – 20 March 2020

**Figure 2.**
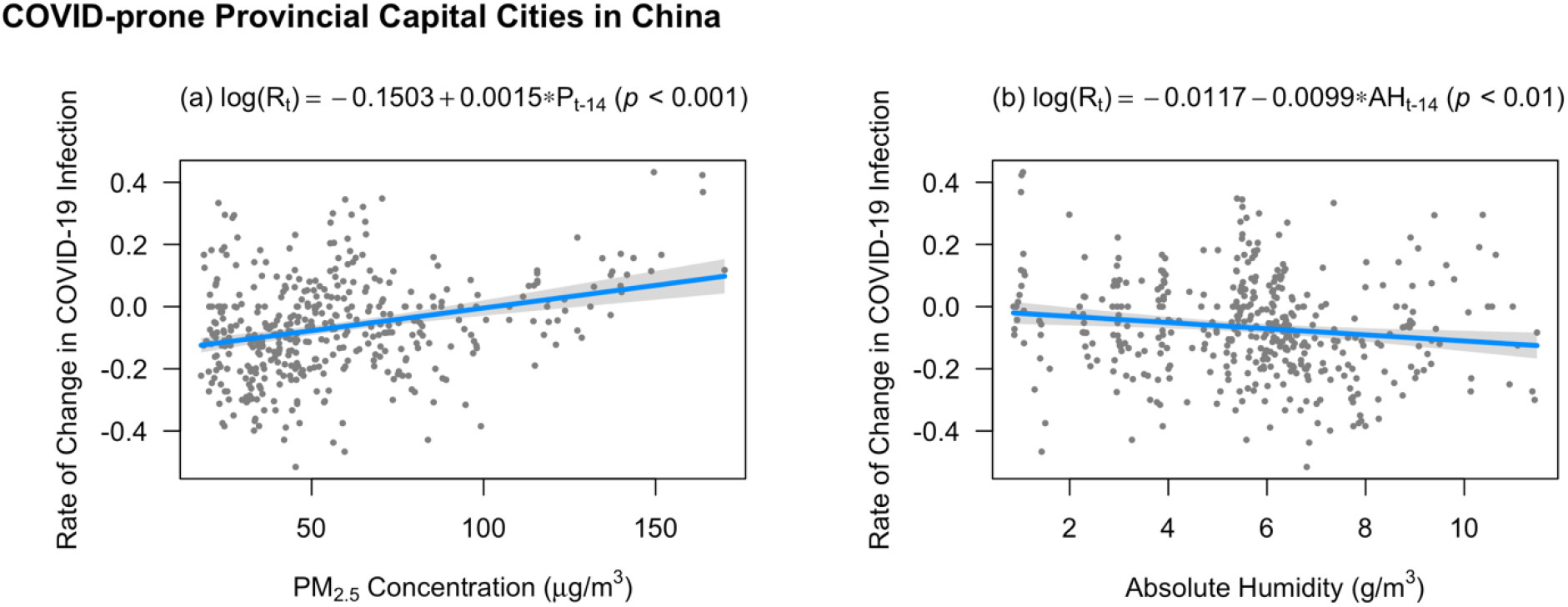
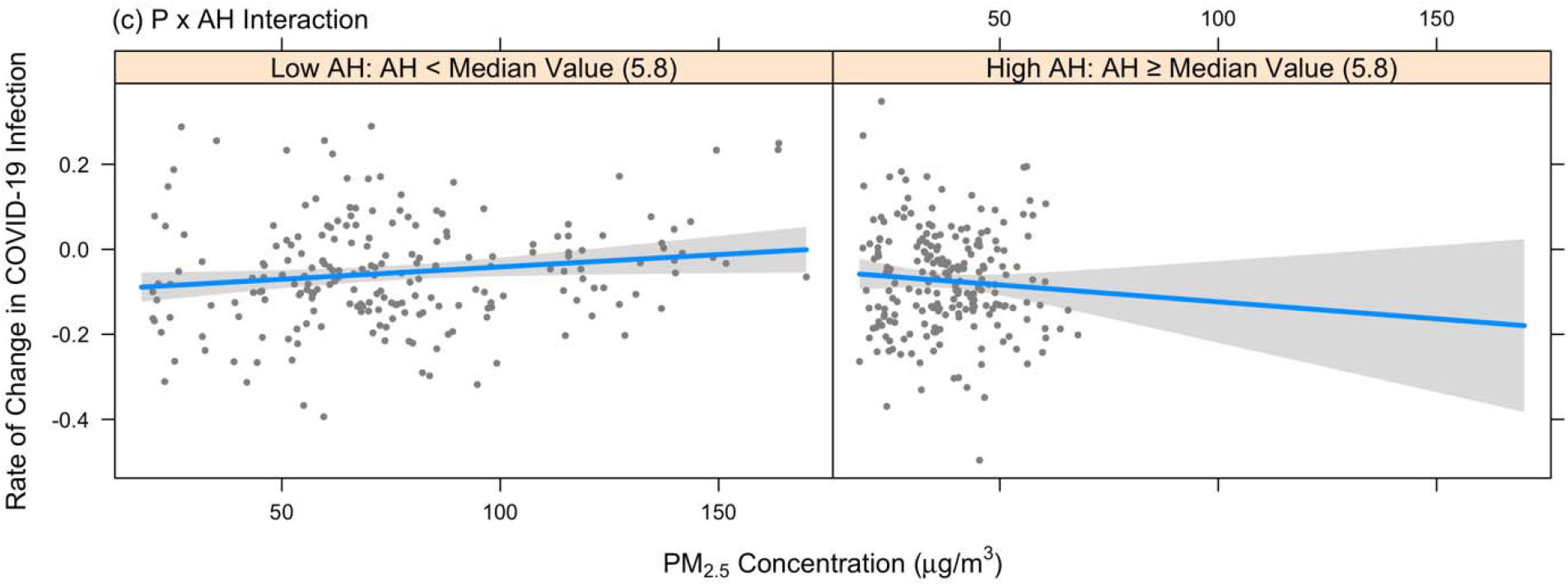
Significant P, AH, and P x AH affecting R across the high infection provincial capital cities in China

### 2.2 Statistical Results

Based on two best-fit regression models (see Section 4.3 for more details), the results of the statistically significant factors that associate with R across the high infection provincial capital cities in China and in Wuhan are shown in Table 2(a) and 2(b), respectively. In order to better illustrate the relationship between P, AH, and R, the individual regression plots of P and AH and their interaction are shown in Figure 2 (high infection provincial capital cities) and Figure 3 (Wuhan only).

**Table 1.** Research objectives and procedures

|  |  |
| --- | --- |
| **Primary Objective** | • Explore the statistical relationship, and determine the causal effects, if any, between daily outdoor P and R across the COVID-prone provincial capital cities in China, including Wuhan, the origin of COVID-19 outbreak.<br>• To achieve this objective, we built two statistical models that can best address the following challenges in statistical analysis:<br>○ Redefinitions and potential delays in infection case reporting<br>○ Incubation period |
|  | ○ Confounders and confounding biases<br>○ Collinearity<br>○ Non-linearity<br>○ Interaction between P and meteorology |
| **Secondary Objective** | • Highlight the conditions under which R can be reduced, and effective public health measures that can be employed to facilitate this.<br>• Add weight to the current observations that COVID-19 can be airborne and that particulates can be carriers of the viral droplets. |

**Table 2.** Statistically significant factors that associate with R (a) across all the high infection provincial capital cities in China and (b) in Wuhan (from 1 January to 20 March 2020).

| **(a) High Infection Provincial Capital Cities in China** |  |  |
| --- | --- | --- |
| **Dependent Variable:** $R_t$ | | **Number of Observations:** $n=421$ |
| **Number of Independent Variables:** 6 |  | **Adjusted $R^2$:** 39.6% |
| **Independent Variables** | **Coefficient with 95% CI** | **p-value** |
| Intercept | -0.0873 [-0.2307, 0.0562] | 0.2324 |
| $R_{t-1}$ | 0.2814 [0.2012, 0.3616] | $2.00 \times 10^{-11}$ *** |
| $NM_{t-L}$ | 0.2284 [0.0721, 0.3846] | 0.0043 ** |
| $P_{t-L}$ | 0.0016 [0.0007, 0.0025] | 0.0006 *** |
| $AH_{t-L}$ | 0.0152 [0.0038, 0.0265] | 0.0089 ** |
| $T_t$ | -0.0060 [-0.0074, -0.0045] | $1.33 \times 10^{-14}$ *** |
| $P_{t-L} \times AH_{t-L}$ | -0.0002 [-0.0004, -0.0000] | 0.0170 * |
| **(b) Wuhan Only** |  |  |

| **Dependent Variable: $R_t$** |  | **Number of Observations: $n=49$** |
| --- | --- | --- |
| **Number of Independent Variables: 3** |  | **Adjusted $R^2$: 67.7%** |
| **Independent Variables** | **Coefficient with 95% CI** | **$p$-value** |
| Intercept | -0.2466 [-4.2653, -0.6673] | 0.0008 ** |
| $P_{t-L}$ | 0.0856 [0.0400, 0.1311] | 0.0005 *** |
| $AH_{t-L}$ | 0.3849 [0.0938, 0.6759] | 0.0107 * |
| $P_{t-L} \times AH_{t-L}$ | -0.0140 [-0.0217, -0.0063] | 0.0007 *** |
| **Notes**<br>1. For (a) High infection provincial capital cities in China, P, AH, and NM were lagged and averaged by $L=14$ days. For (b) Wuhan, P and AH were lagged and averaged by $L=14$ days.<br>2. For (b) Wuhan, D and CM were not included in the initial stepwise regression model since they were constant within the city.<br>3. * $p$-value < 0.05, ** $p$-value < 0.01, *** $p$-value < 0.001 | | |

**Figure 3.**
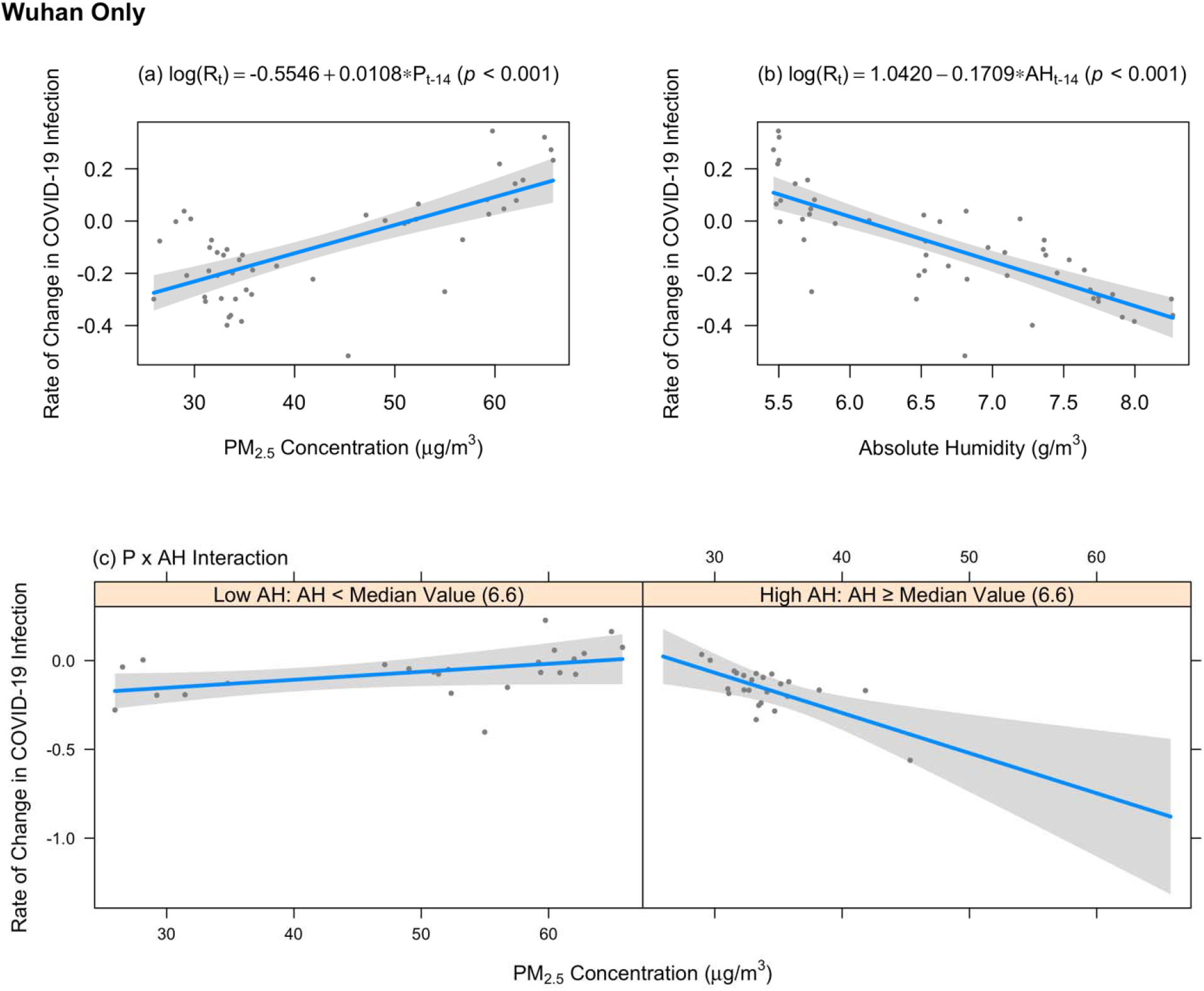
Significant P, AH, and P x AH affecting R in Wuhan

As shown in Table 2(a), P, AH, NM, and T were significant factors affecting R across the high infection provincial capital cities in China. A higher P was associated with a higher R in China. When only looking at the effect of P on R, a 10 µg/m^3^ increase in P was associated with a 1.5% increase in R (*p* < 0.001; see Figure 2(a)). Moreover, AH was a significant factor for accounting R in China (*p* < 0.01). As shown in Figure 2(b), when only looking at the effect of AH on R, a higher AH decreased R. NM and T were also significant factors for predicting R in China. NM had a significant statistical correlation with R, and an increase in R was observed along with the increase in NM (*p* < 0.01). T had a significant statistical correlation with R, and a decrease in R was observed along with the increase in T (*p* < 0.001). The best-fit day-lag for P, AH, and NM was fourteen. Furthermore, based on the significant covariates identified in Table 2(a), the causal effect of P on R was established via matching, by addressing the confounding biases. The result was consistent with our main findings. On average, across the high infection provincial capital cities, days with higher P (≥ 100 µg/m^3^) resulted in a 16.7% increase in R compared to days with lower P (< 100 µg/m^3^), after controlling for the important confounding factors including AH, NM, and T (see Table S6 in Appendix, p 6).

Moreover, the interaction between P and AH was significant across the high infection provincial capital cities in China (see Table 2(a)). To further investigate the interaction between P and AH, AH was categorized into two levels according to its median value. When AH is less than 5.8 g/m^3^, the median of AH values, a higher P and AH gave a higher R. When AH > 5.8 g/m^3^, a higher P and AH resulted in a lower R. As shown in the left part of Figure 2(c), when a higher P interacted with a lower AH, a higher R was still observed on average. In contrast, as shown in the right part of Figure 2(c), the effect of a higher P on R (in increasing trend) was counter-balanced by the effect of a higher AH on R (in decreasing trend).

Furthermore, when Wuhan, the city with the greatest number of confirmed cases during the period of study, was included in our analysis, similar statistical results were observed. As shown in Table 2(b) and Figure 3, the effects of P and AH on R in Wuhan were also statistically significant (*p* < 0.05). However, NM and T were not statistically associated with R in Wuhan (*p* > 0.05). Compared to all cities in China, the effect of P on R in Wuhan was higher. When only looking at the effect of P on R, a 10 µg/m^3^ increase in P was associated with a 10.8% increase in R in Wuhan (*p* < 0.001; see Figure 3(a)). Moreover, the interaction effect of P and AH on R in Wuhan was statistically significant (*p* < 0.001). As shown in Figure 3(c), when a higher P interacted with a lower AH, a higher R was observed on average, but when AH was higher, the effect of a higher P on R was counteracted by the effect of AH on R.

## 3. Discussion

Recent COVID-19 studies have investigated whether D, CM, meteorology, and lockdown are associated with significant statistical effects on viral infection. Some studies ascertained that meteorological effects demonstrate an association with COVID-19 transmission.^2^ Earlier influenza studies have suggested that exposure to P with and without interactions with meteorological effects might increase the risks of influenza infection. Earlier epidemiological studies have also identified that API (measuring PM_10_ as the primary pollutant) and meteorological effects are associated with SARS/MERS. In the US and Europe, long-term exposures to P and to NO_2_ have been reported as the predictors of COVID-19 mortality. Recently, increasing evidence in China and Italy argued that air pollution was a significant attributor to COVID-19 infection. Previous studies conducted in China have concluded that P is associated with COVID-19 infection, though it does not comprehensively account for the change in TC and the inadequacy in COVID-19 confirmed case definition, and the confounding effect of D and CM. Recent scientific studies have also pointed towards the significant potential of airborne transmission by COVID-19.^22^

To identify whether P affects R across 18 high infection provincial cities in China, including Wuhan, our regression model takes into account all high potential confounders, including meteorological variables, NM, D at the provincial or city level, and CM at the provincial level, including eight major diseases that can potentially decrease immunities and increase the risk of COVID-19 infection.^23,24^ In addition, the time-lag effect on P, meteorology, and NM, were addressed.

Our model outperforms other existing air-pollution related COVID-19 epidemiological studies in four ways. First, instead of observing the absolute number of infections, which can be inadequate due to possible human or systemic deficiency, in relation to testing methods and changes in case definition, our study examines R, the rate of change in COVID-19 infection (see Section 4.1). R can sufficiently reflect the relative change in infection numbers, provided that the adjusted COVID-19 infection trends are consistent. Therefore, our methodology is capable of providing greater insights than the previous epidemiological studies on air pollution and COVID-19 infection/mortality, which are focussed on the absolute number of infections^13,14^ instead of infection rate^25^, or R.

Second, our study has addressed a wide spectrum of possible confounders that may affect observations concerning the effect of P on R, including key meteorological, NM, D, and CM variables. This stands in contrast to existing work that has explored the effect of air pollution on COVID-19 infection/mortality by controlling for only the meteorological variables,^13,26^ or controlling for the meteorological variables and simple D variables without considering the lockdown and the CM confounders.^27^ Using stepwise regression, results have shown that AH was a significant predictor of R in the high infection provincial capital cities in China. In addition, NM and T were significant predictors of R in China, suggesting that an increase in the net move-in mobility could increase R and the unobserved time-varying effects such as a decrease in mobility within a city due to lockdown could reduce R. Further, our work has in parallel sought to address issues of non-linearity, collinearity, and time-lag (see Section 4.2). This is particularly critical for precision modelling when (1) the statistical relationships between meteorology and R could be non-linear, (2) some covariates among the meteorology, demographics, or co-morbidity variables are often collinear, and (3) the short-term effects of P, meteorology, and NM on R could be lagged due to incubation period. By accounting for the non-linearity, collinearity, and time-lag between some of our confounders and R, our model provides a more reliable and rigorous scientific prediction concerning how and when P will affect R across the high-infection provincial capital cities in China, as well as in Wuhan, in contrast to some prior air pollution-related COVID-19 infection/mortality models which have yet taken non-linearity or other statistical characteristics into account.^7,27^

Third, this is the first study that pursues the individual effects of P and AH on R, as well as the interaction effect of P and AH on R, covering (1) the high infection provincial capital cities in China and (2) Wuhan only. Our study based on (1) ascertained that a higher P increased R, and a higher AH decreased R. A 10 µg/m^3^ increase in P was associated with a 1.5% increase in R in China on average (*p* < 0.001; see Figure 2(a)). Further, when P interacted with AH, their interaction effect on R was significantly negative (Coefficient = −0.0002, *p* < 0.05, see Table 2(a)). When breaking down AH into two groups according to its median value, if AH ≤ 5.8 g/m^3^, a higher P still led to a higher R (see Figure 2(c)). However, if AH > 5.8 g/m^3^, the effect of a higher P on R was counteracted by the effect of AH on R. For (2), similar results can be observed. However, P had a much stronger positive effect on R than in (1). A 10 µg/m^3^ increase in P was associated with a 10.8% increase in R in Wuhan on average (*p* < 0.001; see Figure 3(a)). A statistically significant interaction effect between P and AH was also observed in Wuhan (Coefficient = −0.0140, *p* < 0.001, see Table 2(b)). A higher P led to a higher R when AH is lower, but the effect of a higher P on R was counteracted by the effect of a higher AH on R (see Figure 3(c)).

Finally, to the best of our understanding, this is the first international study that claimed a causal relationship between P and R across the high infection Chinese provincial capital cities, via matching. Each high P exposure day was matched with a low P exposure day sharing similar background covariates such as AH and NM to estimate the causal effect (Appendix p 6). This causal relationship between immediate P exposure and R (i.e. a higher P can increase R, see Table 2 and Table S6 in Appendix p 6), when combined with the recent reports that particulates less than 10 µm in size could facilitate the deposition of COVID-19 viral droplets and be suspended in the air,^17^ further substantiates the recent observations regarding the risks of airborne infection.^18,20^

A limitation of this study is that the statistical relationship between P and R in (1) (Coefficient = 0.0015, *p* < 0.001; see Figure 2(a)) was less strong as compared to (2) (Coefficient = 0.0108, *p* < 0.001; see Figure 3(a)). This might be due to the lack of data points at higher P and AH in (1) (see Figure 2), though P, AH, or P × AH was the statistically significant predictor of R across the high infection provincial capital cities in China and in Wuhan only (*p* < 0.05, see Table 2).

Our findings call for immediate medical and public health attention concerning the significance of P in exacerbating R. Controlling and reducing outdoor P, and reducing the possibility for outdoor P to be used as a carrier for COVID-19 viruses, have never been as urgent as they are now. Public health measures such as installing air purifiers, both indoors and outdoors, can help reduce P and alleviate the situation.^28^ Alternatively, improving air ventilation, both indoors and outdoors, presents another possibility.^29^ Despite previous claims that TEMP and WS will temper the COVID-19 infection,^30,31^ our result showed no statistical significance for these factors. Nevertheless, our study supports recent findings on AH that a lower AH may increase COVID-19 infection.^2^ Our study further investigates the interaction of P and AH, and suggests that the effect of a higher P on R was counteracted by the effect of a higher AH on R. Moreover, the possibility for airborne infection of COVID-19 is too high a cost to be ignored and proper public health measures, such as requiring citizens to wear face masks, should be established to reduce the possibility of COVID-19 infection through air, especially for countries with high population densities and mobilities, and high particulate pollution. Given that the best fit model was obtained using a 14-day time-lag, this implies that on average, it would take 14 days for the significant factors, including P, AH, and NM, to have an influence on a COVID-19 patient and the patient to become symptomatic. This will have important public health implications on the number of days needed for quarantine for effective COVID-19 detection.

## 4. Method

### 4.1 Data Collection and Procedure

We collected data covering the daily P and the daily number of confirmed infections of 31 provincial capital cities in China, covering the period from 1 January to 20 March 2020 (see Figure S1 in Appendix p 3). This was the period when COVID-19 infection was first officially announced in China, the lockdown measures were strictly exercised in Wuhan and other parts of China, and the number of confirmed cases peaked and dropped (see Figure S1 in Appendix p 3). Other data at the provincial city-level were also collected on a daily basis (including meteorology and NM) or on a yearly basis (including D and CM) from internet sources and official statistical documents (see Table S1 in Appendix p 2). A full description of the dependent variable and the independent variables adopted for our statistical modelling is listed in Appendix (p 3-5). Table 1 highlights our research objectives and procedures.

### 4.2 Data Pre-processing

Earlier COVID-19 studies expressed reservations concerning the number of infection cases reported, given inadequate testing capacity, the change in confirmed case definition, and undiscovered and undocumented asymptomatic cases.^3,32,33^ In order to address the delay in testing capacity and the change in case definition and their effects on reported cases, we used R, rate of change, as the dependent variable, in order to capture the relative change in COVID-19 infection during the study period. By using R, even if the number of reported infections might deviate, the relative change in infection could still be accounted for, provided that the reporting trends remain consistent.

Moreover, to obtain consistent reporting trends of COVID-19, a four-step data adjustment method was used to reduce the noise in the collected daily confirmed COVID-19 infection data. First, to make valid statistical conclusions, 13 cities with a cumulative number of confirmed cases less than 50 were removed due to the small sample size. This cut-off value was based the assumption that at least five types of independent variables are of our interest (including P, meteorology, NM, D, and CM) and the rule of thumb that each independent variable requires at least ten samples. The remaining eighteen cities were considered the COVID-prone provincial capital cities. Second, for each city, to address the potential delay between the onset and the confirmation of COVID-19 infection, the adjusted daily confirmed COVID-19 infection cases were calculated by a rolling window of the observed daily confirmed cases reported in the following *W* days (including the current day). The rolling window is a simple interpolation technique that smoothened out the short-term fluctuations in the city-specific epidemic curve, while allowing for the backfill of delayed confirmed cases. More specifically, the adjusted number of confirmed cases on day *t* was calculated as the average of the number of confirmed cases reported from day *t* to *t + W* − 1 (see Eq(1)).

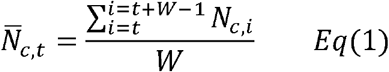

where *N*_*c,t*_ denotes the number of confirmed cases reported on day *t* in city *c. W* was set to 7 to address the reporting delay in COVID-19 case confirmation (which was estimated to be 7 days to 10 days^33^) and to account for the day-of-week fluctuations in case reporting. Then, after the adjustment of daily confirmed cases, for each day in each city, data points with no COVID-19 cases were removed, given that the spread of COVID-19 infection already stopped. Finally, for each selected city, daily R values were calculated throughout the study period (see Eq(2)).

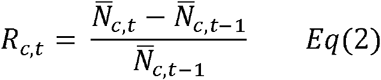

where 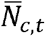 denotes the number of adjusted confirmed cases reported on day *t* in city *c*. For all R values across the selected cities, the mean and standard deviation of R were calculated. R values out of the normal range (mean ± three times standard deviation) were considered outliers and were removed.

### 4.3 Statistical Analysis

Using stepwise regression, a final multiple linear regression model including only the statistically significant variables was constructed to model the relationship between daily outdoor P and daily R across the high infection provincial capital cities in China (see Eq(3)), and in Wuhan, China (see Eq(4)), while taking into account potential confounders and interaction terms (see Table S5 in Appendix p. 5), in order to determine if any causal relationship exists between the two variables and any significant interaction between air pollution and meteorology variables (see Table 2(a)-2(b)).

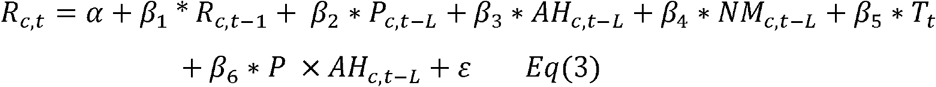

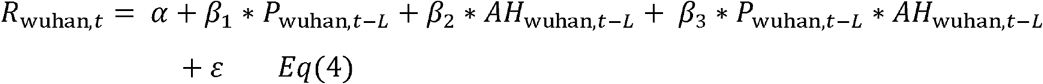

Where *α* is the intercept, the subscript *c* denotes a city, subscript *t* denotes a day, subscript *L* denotes the time lag for P, AH, and NM, and E serves as the error term. *L* ranges from one to fourteen days. R denotes the rate of change in the daily number of confirmed COVID-19 infections. A one-day time-lag variable representing the R of the previous day was also included in the model for China (the high infection provincial capital cities) as an autoregressive term to account for the temporal auto-correlation among R time-series. P denotes the PM_2.5_ concentration. NM denotes the net move-in mobility. AH denotes the absolute humidity. T is a variable representing the number of days since 1 January 2020, reflecting the time trend during the period of study. Two-sided *p*-values < 0.05 were considered significant for the statistical analysis.

Due to the lengthy asymptomatic incubation period before the onset of COVID-19 symptoms, the corresponding time-lag in P, meteorology, and NM was accounted for by our statistical model for both China and Wuhan, using the multi-day average lag model, based on previous air-pollution related epidemiological studies.^6^ We determined the best fit lag-time from day 1 to day 14, with the assumption that the mean incubation period could cover a maximum of 14 days.^33^

To estimate the causal effect of P on R, our model for China and Wuhan had to cover the potential confounders. Independent variables, including meteorology (AH, temperature (TEMP), air pressure (AP), and wind speed (WS)), and NM, were covered in the statistical analysis as the confounders. Moreover, D (population density, age, sex, income, GDP per capita, and education) and CM (high blood pressure, diabetes, chronic obstructive pulmonary disease (COPD), stroke, obesity, asthma, Alzheimer’s disease (AD), and HIV/AIDS) were included in the statistical analysis to control for the provincial/city-level fixed effects. T and day of week were included in the statistical analysis to control for the time-varying fixed effects and the recurrent fixed effects. The statistically significant factors were kept in the final fitted regression model. Furthermore, matching was adopted to further reduce the confounding biases, by matching a high P day with a low P day, based on the similarities of corresponding confounders, thereby helping one more accurately estimate the causal relationship between P and R in China (see Appendix p 6).

To account for the collinearity in our model for China and Wuhan before the stepwise regression analysis, a Spearman correlation analysis was conducted to select meteorological, D, and CM variables that present low collinearity. First, AH and WS were selected as the meteorological variables for the stepwise regression analysis. We tested the collinearity between TEMP, AP, WS, and AH, and removed TEMP and AP, due to their high collinearity with AH, which was able to account for the transmission of a flu virus, and hence may also be used to account for R^34^ (|Spearman coefficient| > 0.5; see Table S2 in Appendix p 3). Moreover, population density, age (0-14 years old), age (> 65 years old), sex ratio (female/male), and GDP per capita were selected as the D variables for the stepwise regression analysis. We tested the collinearity between D variables, population density, age (0-14 years old), age (> 65 years old), sex ratio (male/female), urban disposable income, GDP per capita, and education level (below high school). We found that all D variables, except for sex ratio and GDP per capita, correlated highly with population density and age. Population density and age might better account for R because (1) population density could account for the close-contact transmission of COVID-19 and (2) an old age could be linked to a lower immune function. Therefore, urban disposable income and education level were removed, due to their high collinearity with population density and age (|Spearman coefficient| > 0.5; Table S3 in Appendix p 4). Furthermore, high blood pressure, COPD, stroke, and asthma were selected as the CM variables for the stepwise regression analysis. We tested the collinearity between CM variables, high blood pressure, diabetes, COPD, stroke, obesity, asthma, AD, and HIV/AIDS. We found that all CM variables, except for stroke and asthma, correlated highly with high blood pressure and COPD, which were more common CM identified from recent COVID infection cases, and might account for R.^21^ Therefore, diabetes, obesity, AD, and HIV/AIDS were removed, due to their high collinearity with high blood pressure and COPD (|Spearman coefficient| > 0.5; Table S4 in Appendix p 4).

To account for the non-linear relationship between the meteorological variables and R in our model for China and Wuhan, a non-linear transformation was applied to the selected meteorological variables, including AH and WS. Two transformation functions, a second order polynomial function and a natural spline function with two degrees of freedom, were attempted to address non-linearity, based on the goodness of fit. From this it was determined that the first-order polynomial function already provided the best fit and was adopted to account for the relationship between AH/WS with R.

Finally, in addition to the investigation of the non-linear effects of meteorological variables on R, to examine whether there was any interaction between P and meteorology on R across the high-infection provincial capital cities in China and in Wuhan during the infection cycle, an interaction term between P and AH was developed for China and Wuhan (Eq (3-4)), based on the statistically significant interaction terms that associate with R (see Table 2).

## Data Availability

The datasets are available from the corresponding authors on reasonable request.

## Data Availability

The dataset used in this study will be made available upon request to the corresponding authors.

## Code Availability

The data processing and statistical analysis code for this study will be made available upon request to the corresponding authors.

## Contributor

JCL and VOL were responsible for conceptualization and initial framework development. YH collected the statistical data. JCL, VOL and YH developed the methodology. YH processed the data and conducted the statistical analysis. YH, JCL and VOL interpreted the results and wrote the full manuscript. JC, JF, JD, and IG provided valuable suggestions and comments on the data input and analysis, and the methodology. JD edited the manuscript. JCL and VOL applied for funding. YH, JCL and VOL contributed equally.

## Acknowledgement

This research is supported in part by the Theme-based Research Scheme of the Research Grants Council of Hong Kong, under Grant No. T41-709/17-N. We acknowledge the data collection by Peiyang Guo, Qi Zhang, and Andong Wang, comments and suggestions from Shan Shan Wang, Zafar Gilani, as well as comments from Joseph Hui and K.W. Wu regarding the non-linear relationship between the independent variables and the dependent variable.

